# Validation of a novel, low-cost, portable MRI-compatible exercise device in healthy volunteers and patients with pulmonary hypertension

**DOI:** 10.1101/2024.07.20.24310708

**Authors:** Ruta Virsinskaite, James T. Brown, Tushar Kotecha, Darren Bower, Jennifer A. Steeden, Javier Montalt-Tordera, Olivier Jaubert, Marianna Fontana, J. Gerry Coghlan, Daniel S. Knight, Vivek Muthurangu

## Abstract

**Introduction:** The value of exercise cardiovascular magnetic resonance (CMR) has been shown in many clinical scenarios. We have developed a MR-compatible exercise apparatus and aim to validate it against the reference standard MR-conventional ergometer.

**Methods:** The novel device consisted of two half-pipes fixed to a wooden base, with participants wearing knee-length socks with a 0.5kg weight in each sock. Increased workload was achieved by increasing the rate of alternating leg flexion and extension in time with a bleep sound of increasing frequency.

Twenty subjects (10 healthy volunteers, 10 patients with pulmonary hypertension) performed two CMR-augmented cardiopulmonary exercise tests (CMR-CPET) using the novel exercise apparatus and a conventional ergometer in a randomised order.

**Results:** Comparing peak metrics elicited on both exercise devices, there was a moderate correlation in peak oxygen consumption (VO_2_, r=0.86, P<0.001), cardiac output (CO, r=0.66, P=0.002), stroke volume (SV, r=0.75, P<0.001), peak heart rate (HR, r=0.65, P=0.002) and peak arteriovenous oxygen content gradient (ΔavO_2_, r=0.71, P<0.001). However, all metrics (except peak SV) were significantly lower from the novel device. Both devices were able to elicit statistically significant differences in VO_2_, HR and RVEF between patients and healthy subjects (P≤0.036).

**Conclusions:** We have created a simple, easy to use and affordable exercise apparatus for CMR environment. This may encourage greater dissemination of exercise CMR in clinical and research practice.

## Background

Exercise Cardiovascular Magnetic Resonance (CMR) is increasingly used to evaluate cardiovascular reserve. Several studies have shown that exercise CMR can be used to unmask underlying cardiac dysfunction, as well as to investigate the causes of exercise intolerance (1). This is particularly true in pulmonary arterial hypertension (PAH) where exercise CMR has been used to assess contractile reserve (2, 3). However, exercise CMR has been slow to enter routine clinical practice due to three main factors: i) excessive motion/breathing during exercise imaging, ii) the cost of MR-compatible exercise equipment, and iii) the complexity of set-up and performing exercise CMR protocols.

Segmented k-space imaging is difficult to perform during exercise because of bulk motion and poor breath-holding. Consequently, most exercise CMR protocols leverage real-time CMR, which is less sensitive to motion and can be acquired during free breathing. Recently, accelerated real-time CMR with sophisticated reconstruction (e.g. compressed sensing) has improved spatio-temporal resolution, which is vital at high heart rates (4, 5). Furthermore, deep learning (DL) has been used to both reconstruct and process real-time flow CMR data during exercise, further simplifying imaging during exercise (6, 7).

Thus, CMR during exercise is becoming easier which should aid clinical translation. However, broader adoption of exercise CMR is still limited by the cost (∼$40K) and the cumbersome nature of commercial MR-compatible ergometers. Several studies have successfully demonstrated ‘in-house’ ergometers (8, 9), but most are complex and would be difficult to manufacture in other centers. In addition, they are not necessarily easier to use than commercial ergometers and this is a significant limitation. Therefore, we have developed a portable, low-cost, and easy to use MR-compatible exercise device that can be easily reproduced/manufactured in any institution. The aim of this study was to compare our novel device against a commercial MR-conditional ergometer in both healthy subjects and PAH patients, using our previously published CMR-augmented cardio-pulmonary exercise testing (CMR-CPET) protocol (2, 10).

## Methods

### Novel Exercise Device

The novel MR-compatible exercise apparatus (Figure 1A) consisted of two 83.3 cm length and 11.2 cm width half-pipes (made from commercially available plastic roof guttering) fixed to a wooden base (using wooden brackets and adhesive without any ferromagnetic components). The dimensions of device necessary for manufacture are shown in Supplementary Figure 1. The wooden base was designed to fit into the recess of the scanner table for stability, replacing the usual cushion (Supplementary Figure 1). Subjects wore knee-length thrombo-embolic deterrent (TED) stockings with a 0.5 kg sandbag placed in the front of each sock. The TED stockings were used to ensure low friction movement, as well as to hold the weight in place, with 0.5kg being chosen empirically after preliminary testing in healthy volunteers. Exercise was achieved by alternating leg flexion and extension in time with a bleep sound that increased in frequency every minute from 60bpm to a plateau of 180bpm. Thus, work was increased through increasing the frequency of motion rather than increasing load. A video demonstrating the exercise technique is included in supplementary material (Supplementary Video 1), as is the 18-minute sound file containing the ramped bleep sounds (Supplementary Audio).

**Figure 1.**
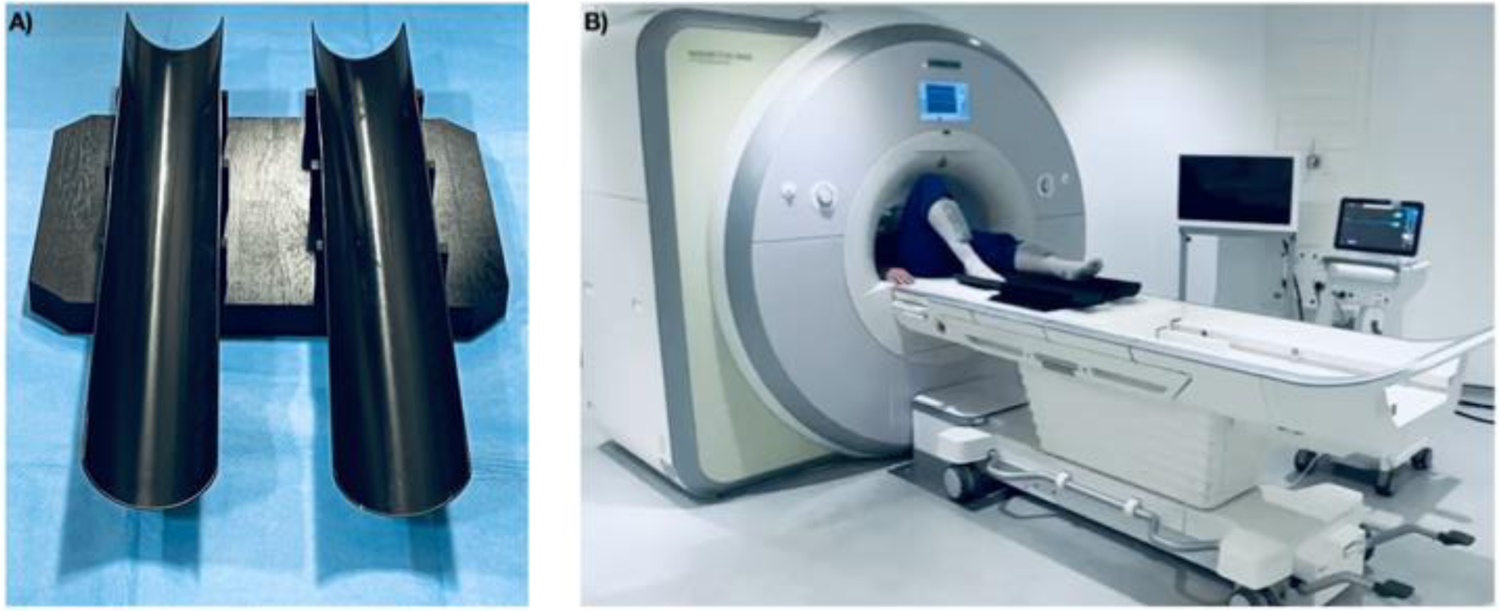
A, The novel exercise apparatus. B, Subject exercising using the novel exercise device.

### Patient population

Ten healthy volunteers and ten patients with PAH were recruited between November 2022 and June 2023. Patients were recruited from the Royal Free Hospital pulmonary hypertension service. Diagnoses were as follows: connective tissue disease associated PAH = 7, porto-pulmonary hypertension = 2, idiopathic PAH = 1. Inclusion criteria were: i) clinical diagnosis of pulmonary hypertension confirmed by right-heart catheterization for patient groups and ii) age 18–80 years. Exclusion criteria were: i) general contraindications to CMR scanning, ii) contraindications to performing exercise test (unstable symptoms, including angina, exertional syncope, WHO class IV symptoms and musculoskeletal disease preventing exercise) and, iii) changes in targeted PAH therapy within three months or during the experimental period. The study was approved by UK National Health Service, Health Research Authority, Research Ethics Committee and the study conformed to the declaration of Helsinki (IRAS project ID 226101; REC reference 17/LO/1499, National Health Service Health Research Authority UK CRN 058274). All subjects provided written informed consent.

### Comparing exercise devices

Exercise CMR studies were performed using both our novel device and a conventional supine bike ergometer on all subjects. The two devices were tested on separate days in a randomised order with a maximum time between tests of six weeks.

The conventional MR-compatible exercise study was performed on a commercially available supine ergometer (Lode BV, Groningen, The Netherlands) and followed a ramped protocol of increasing resistance whilst the patient cycled at 60-70 revolutions per minute as previously described (2). For both exercise studies, peak exercise was defined as onset of exhaustion (defined as an inability to maintain cadence or a verbal indication from the subject). The scanning room was temperature controlled. Full resuscitation facilities were available. Each subject’s electrocardiogram (ECG) and oxygen saturation were monitored continuously using the in-built system in the CMR scanner. This system allowed assessment of rate and rhythm but is not suitable for identification of ischemia. All patients had peripheral venous access during testing for use in resuscitation protocols in the event of clinical instability.

### CMR-augmented cardiopulmonary exercise testing

The CMR-CPET protocol performed on both devices included simultaneous continuous breath-by-breath gas exchange analysis, and real-time CMR evaluation of aortic flow and biventricular volumes acquired on the rest and peak exercise.

Imaging was performed on a standard (length 1200 mm, width 489 mm and height 63 mm) 1.5T CMR scanner (Magnetom Aera, Siemens Healthineers, Erlangen, Germany) using two six-element coils (one spinal matrix, one body matrix). Aortic flow was measured using real-time phase-contrast MR (PCMR) using a variable density golden-angle spiral sequence, with low latency deep learning (DL) reconstruction and segmentation (parameters: slice thickness=6mm, velocity encoding = 300 cm/s, temporal resolution = 35ms, spatial resolution = 2.1 × 2.1mm, acquisition time = 10s (6). Assessment of left ventricular (LV) and right ventricular (RV) volumes was performed using 2D multi-slice sorted golden-angle radial real-time balanced steady state free precession (bSSFP) sequence with DL reconstruction (parameters: slice thickness = 8mm, temporal resolution ∼ 31ms, spatial resolution = 1.7 × 1.7mm, no of slices = 12, acquisition time = 24 R-R intervals (7). All real-time imaging was acquired during free breathing at rest and at peak exercise.

Breath-by-breath gas exchange analysis was performed throughout the exercise protocol using a commercial CPET system (Ultima, MedGraphics, St Paul, Minnesota, USA). The analyzer was placed in the CMR control room and attached to the facemask (Hans Rudolph, Kansas City, Kansas, USA) via a set of modified non-ferromagnetic CMR-compatible sampling tubes (umbilicus) with overall length of 1000 cm which were passed through the waveguide. Gas and flow calibrations were performed before each test and at least 30 min after system initiation. All measurements were taken at body temperature and ambient pressure.

### Data analysis

All post-processing of the reconstructed images was performed using ‘in-house’ plug-ins for the open-source OsiriX DICOM software version 9.0.1 (OsiriX Foundation, Geneva, Switzerland) (11). PCMR flow data was automatically segmented as part of reconstruction with manual operator correction if required (6). Stroke volume (SV) was calculated by integrating the flow curve across a single R-R interval. Cardiac output (CO) was given by SV x heart rate (HR). Biventricular endocardial borders were traced manually, excluding papillary muscle and trabeculae from the blood pool, on the short-axis images at end-diastole and end-systole, identified by visual assessment of the largest and smallest cavity areas, respectively. Biventricular stroke volumes were calculated as the difference between the end-diastolic volume (EDV) and end-systolic volume (ESV), and ejection fraction (EF) was determined as (SV/EDV) × 100. All volumetric data and cardiac output were indexed to body surface area (BSA). The VO_2_ and respiratory exchange ratio (RER) measurements were time-registered to CMR data. The VO_2_ was indexed to body weight. Arteriovenous oxygen content gradient was calculated as ΔavO2 = VO2/CO (using non-indexed data).

### Statistical analysis

All statistical analyses were performed using R (version 4.1.2, The R Foundation for Statistical Computing, Venna, Austria). Data were examined for normality using the Shapiro–Wilk normality test. Descriptive statistics were expressed as mean (±standard deviation) for normally distributed data and median (interquartile range) for non-normally distributed data. Agreement between metrics measured whilst undergoing exams on both exercise devices was assessed using Spearman rank correlation coefficient and Bland Altman analysis, with the significance of the bias assessed using a paired t-test. Differences in CMR-CPET metrics between rest and exercise were (for both devices) were assessed using paired t-tests (normal data) and Wilcoxon signed rank tests (non-normal data), while differences between patients and healthy controls were evaluated using t-tests (normal data) and Mann-Whitney tests (non-normal data). Sex distribution between the groups was assessed using the Chi-squared test. A P-value<0.05 was considered statistically significant.

## Results

### Demographics and Resting CMR

Compared to healthy volunteers, patients were older (48±14yrs vs 29±9yrs, P=0.004) and predominantly female (70% vs 40%, P=0.04). The average height for all participants was 1.69±0.09 m (maximum 1.84 m, minimum 1.50 m) with average body mass index of 24.76±3.8 kg/m^2^. There was excellent correlation in left ventricular volumes (LVEDV, r=0.85, P<0.001; LVESV, r=0.82, P<0.001) and right ventricular volumes (RVEDV, r=0.90, P<0.001; RVESV, r=0.83, P<0.001) acquired at rest prior to exercise using both devices (Table 1). There was also good correlation for both resting SV (r=0.52; P=0.02) and CO (r=0.55, P=0.01). There were no significant biases in data acquired at rest, except for SV which was 9% (P=0.01) lower on the novel apparatus.

**Table 1.**
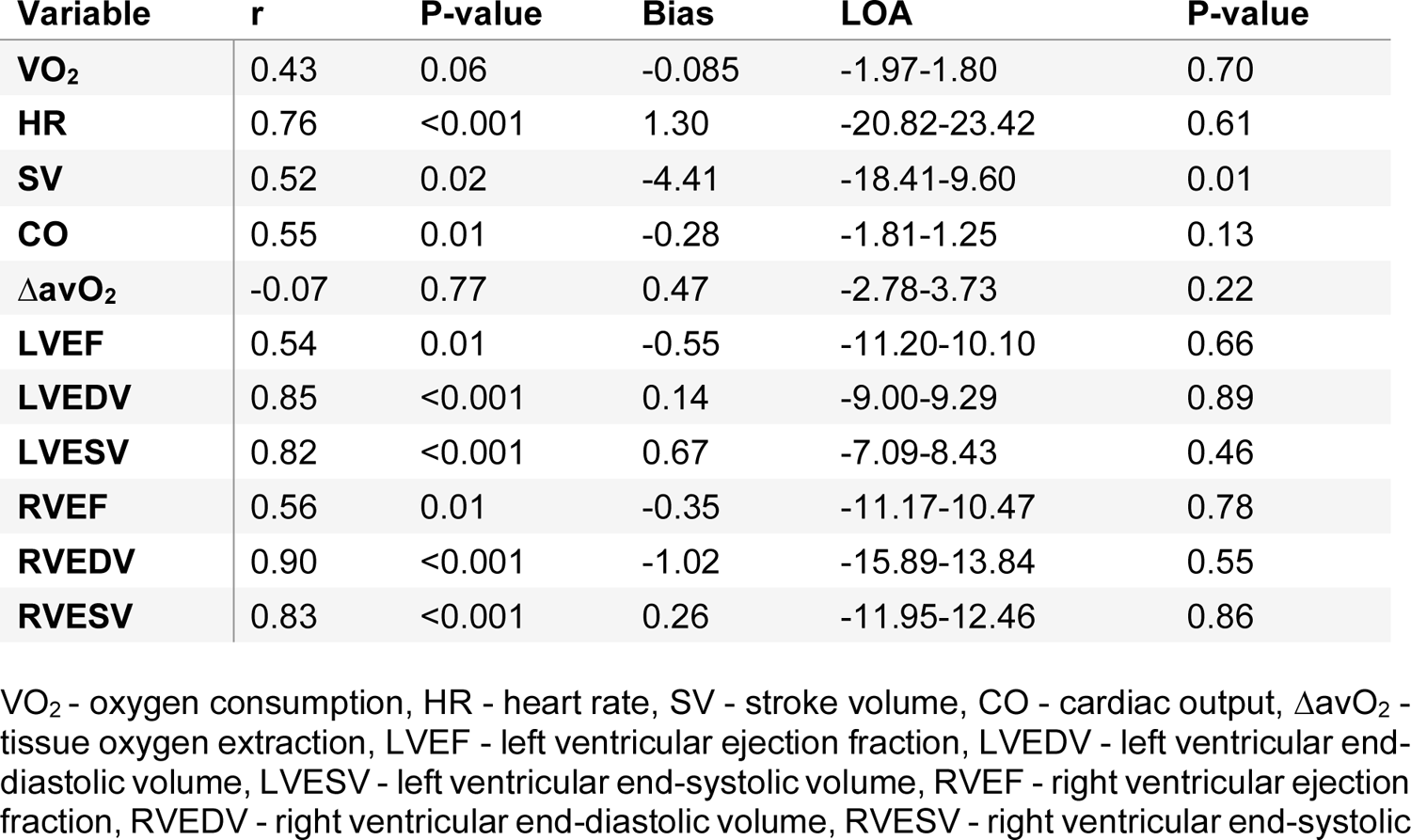
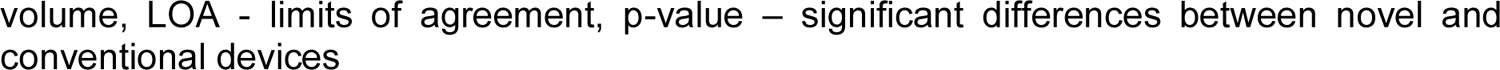
Resting CMR metrics.

| Variable | r | P-value | Bias | LOA | P-value |
| --- | --- | --- | --- | --- | --- |
| VO<sub>2</sub> | 0.43 | 0.06 | -0.085 | -1.97-1.80 | 0.70 |
| HR | 0.76 | <0.001 | 1.30 | -20.82-23.42 | 0.61 |
| SV | 0.52 | 0.02 | -4.41 | -18.41-9.60 | 0.01 |
| CO | 0.55 | 0.01 | -0.28 | -1.81-1.25 | 0.13 |
| ΔavO<sub>2</sub> | -0.07 | 0.77 | 0.47 | -2.78-3.73 | 0.22 |
| LVEF | 0.54 | 0.01 | -0.55 | -11.20-10.10 | 0.66 |
| LVEDV | 0.85 | <0.001 | 0.14 | -9.00-9.29 | 0.89 |
| LVESV | 0.82 | <0.001 | 0.67 | -7.09-8.43 | 0.46 |
| RVEF | 0.56 | 0.01 | -0.35 | -11.17-10.47 | 0.78 |
| RVEDV | 0.90 | <0.001 | -1.02 | -15.89-13.84 | 0.55 |
| RVESV | 0.83 | <0.001 | 0.26 | -11.95-12.46 | 0.86 |

VO<sub>2</sub> - oxygen consumption, HR - heart rate, SV - stroke volume, CO - cardiac output, ΔavO<sub>2</sub> - tissue oxygen extraction, LVEF - left ventricular ejection fraction, LVEDV - left ventricular end-diastolic volume, LVESV - left ventricular end-systolic volume, RVEF - right ventricular ejection fraction, RVEDV - right ventricular end-diastolic volume, RVESV - right ventricular end-systolic
volume, LOA - limits of agreement, p-value – significant differences between novel and conventional devices

### Exercise feasibility

All subjects successfully completed both exercise protocols and no subjects required medical intervention. All subjects (irrespective of height or BMI) were able to perform exercise on the novel device without being their legs being obstructed by the scanner bore. Peak RER≥1.0 was achieved in all participants whilst exercising on the novel exercise apparatus. However, three patients who exercised to exhaustion on the conventional ergometer did not achieve anaerobic threshold (RER<1.0). The exercise time was significantly shorter with the novel exercise device for both healthy controls and patients (heathy control: conventional 15.02±4.15 minutes, novel 10.27±4.75 minutes, P<0.001; patients: conventional 8.35±2.42 minutes, novel 6.65±3.48 minutes, P=0.002).

### Agreement in exercise CMR-CPET metrics

When comparing peak exercise CMR-CPET metrics obtained using both exercise devices (Table 2), there was a moderate correlation in peak VO_2_ (r=0.86, P<0.001, Figure 2A), peak CO (r=0.66, P=0.002, Figure 2B), peak SV (r=0.75, P<0.001, Figure 2C), peak HR (r=0.65, P=0.002) and peak ΔavO_2_ (r=0.71, P<0.001). However, all these metrics (except peak SV) were significantly lower (8-37%, P≤0.04) on the novel device (Table 2, Figure 2D-F). There was also good/moderate correlation in peak RV volumes (r≥0.77, P<0.001) and peak LV volumes (r≥0.66, P≤0.002). Although there was no significant bias in RV volumes between the two devices (Table 2), peak LV volumes were significantly higher using the novel device (between 6-18%, P≤0.04). There was a trend towards no significant correlations in peak RVEF and LVEF (RVEF: r=0.42, P=0.06, Figure 3A and LVEF: r=0.39, P=0.09, Figure 3B) with no significant bias (Figure 3C-D).

**Figure 2.**
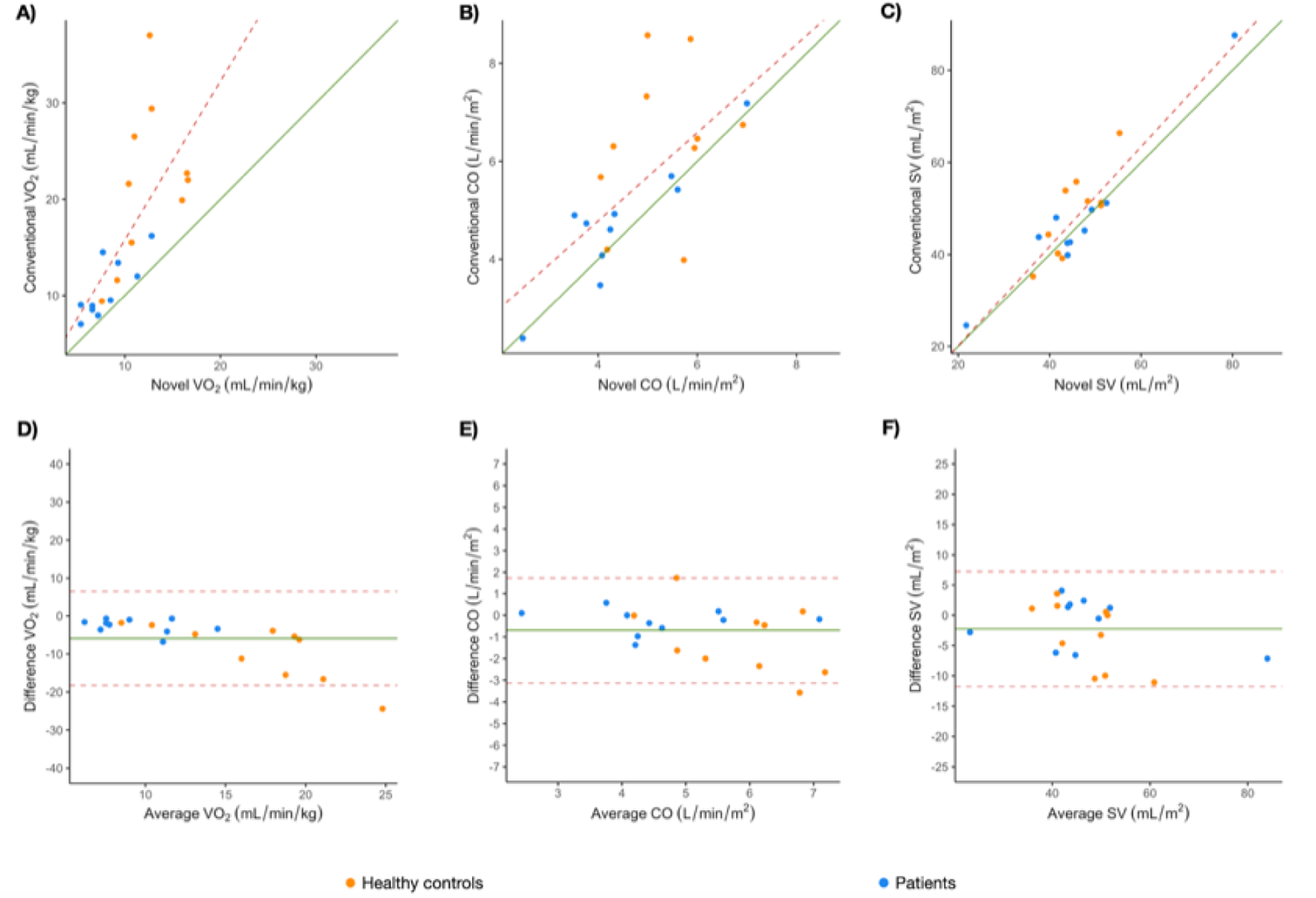
A-C Scatter plots of hemodynamic parameters acquired using the novel and conventional MR-ergometer. A, Oxygen consumption (VO_2_). B, Cardiac output (CO). C, Stroke volume (SV). D-F Bland-Altman charts comparing hemodynamic parameters acquired using the novel and conventional MR-ergometer. A, VO_2_. B, CO. C, SV.

**Figure 3.**
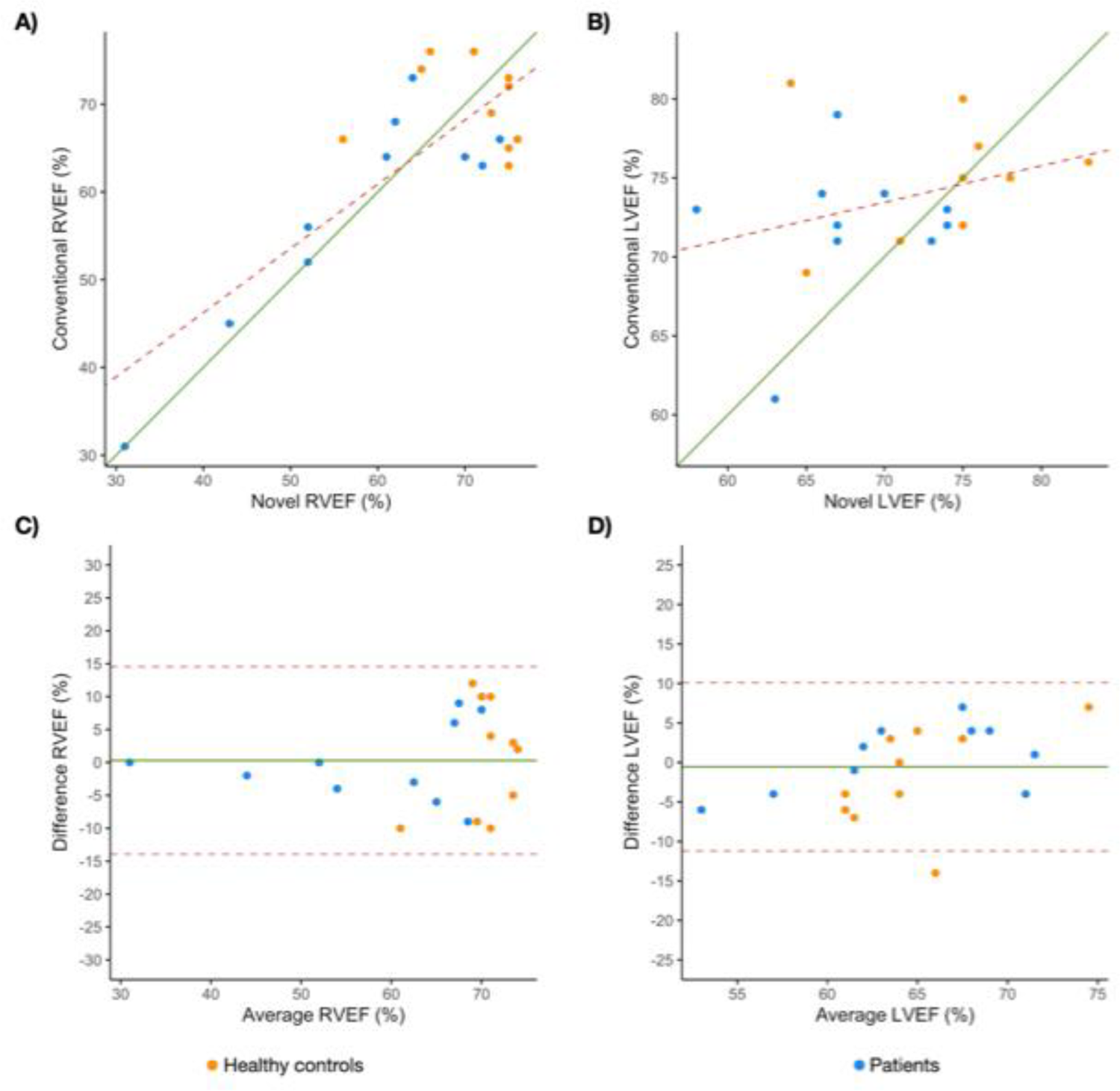
A-B Scatter plots of hemodynamic parameters acquired using the novel and conventional MR-ergometer. A, Right ventricular ejection fraction (RVEF). B, Left ventricular ejection fraction (LVEF). C-D Bland-Altman charts comparing hemodynamic parameters acquired using the novel and conventional MR-ergometer. A, RVEF. B, LVEF.

**Table 2.** Peak CMR metrics.

| Variable | r | P-value | Bias | LOA | P-value |
| --- | --- | --- | --- | --- | --- |
| **VO<sub>2</sub>** | 0.86 | <0.001 | -5.91 | -18.25-6.42 | <0.001 |
| **HR** | 0.65 | 0.002 | -8.85 | -44.89-27.19 | 0.04 |
| **SV** | 0.75 | <0.001 | -2.25 | -11.76-7.26 | 0.05 |
| **CO** | 0.66 | 0.002 | -0.70 | -3.13-1.74 | 0.02 |
| **ΔavO<sub>2</sub>** | 0.71 | <0.001 | -2.53 | -7.72-2.66 | <0.001 |
| **LVEF** | 0.39 | 0.09 | -2.80 | -15.07-9.47 | 0.06 |
| **LVEDV** | 0.75 | <0.001 | 4.40 | -12.95-21.75 | 0.04 |
| **LVESV** | 0.66 | 0.002 | 3.43 | -8.81-15.66 | 0.02 |
| **RVEF** | 0.42 | 0.06 | 0.30 | -13.95-14.55 | 0.86 |
| **RVEDV** | 0.88 | <0.001 | 2.79 | -13.02-18.60 | 0.14 |
| **RVESV** | 0.77 | <0.001 | 0.60 | -14.07-15.27 | 0.73 |

VO<sub>2</sub> - oxygen consumption, HR - heart rate, SV - stroke volume, CO - cardiac output, ΔavO<sub>2</sub> - tissue oxygen extraction, LVEF - left ventricular ejection fraction, LVEDV - left ventricular end-diastolic volume, LVESV - left ventricular end-systolic volume, RVEF - right ventricular ejection fraction, RVEDV - right ventricular end-diastolic volume, RVESV - right ventricular end-systolic volume, LOA - limits of agreement, p-value – significant differences between novel and conventional devices

### Distinguishing Patient from Controls with both protocols

Peak exercise metrics for healthy controls and patients using both exercise devices are shown in Table 3. During exercise VO_2_ and HR significantly increased in both patients and healthy controls on both devices (P<0.001). Patients had significantly lower peak VO_2_ and peak HR than healthy volunteers whilst exercising on the both novel and conventional devices (VO_2_ - conventional P=0.0045; novel P=0.015, Figure 4A; HR - conventional P<0.001; novel P=0.036).

**Figure 4.**
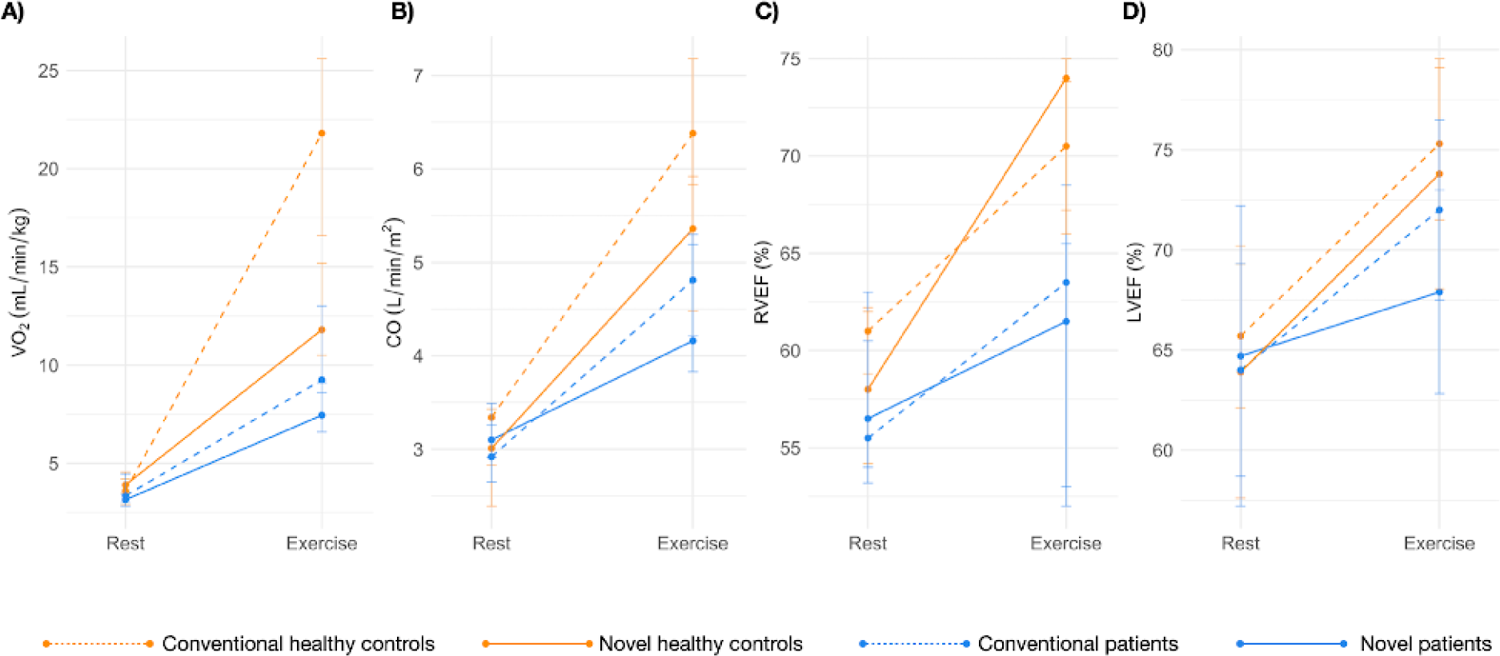
CMR-CPET metrics at rest and peak exercise on both devices for patients and healthy controls. A, Oxygen consumption (VO_2_). B, Cardiac output (CO). C, Right ventricular ejection fraction (RVEF) D, Left ventricular ejection fraction (LVEF).

**Table 3.**
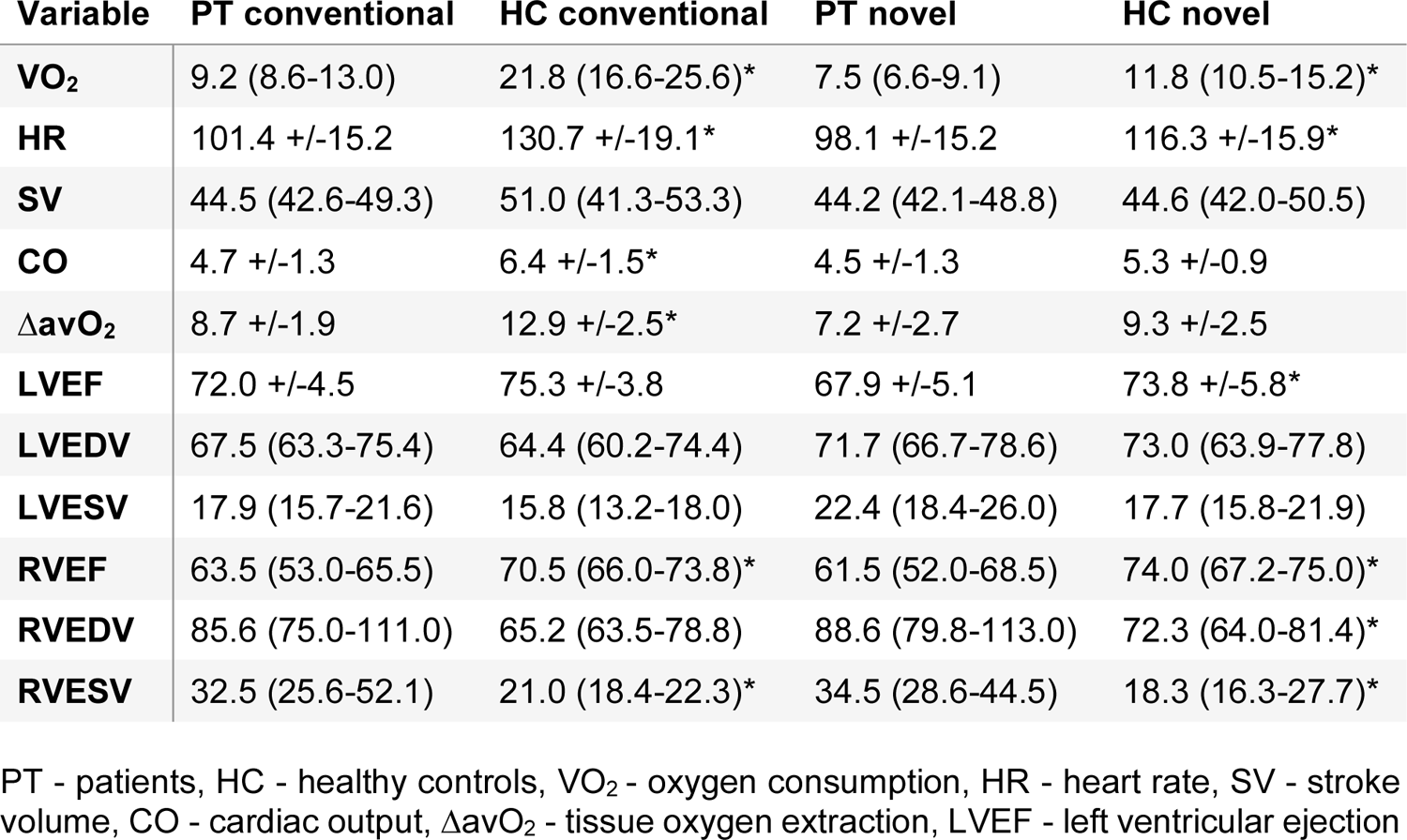
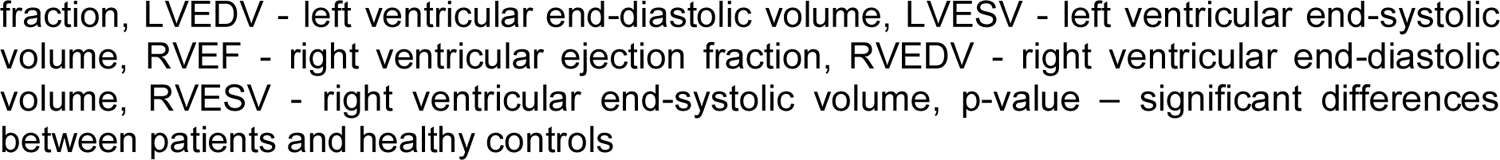
Exercise CMR metrics for novel and conventional exercise devices.

| Variable | PT conventional | HC conventional | PT novel | HC novel |
| --- | --- | --- | --- | --- |
| **VO<sub>2</sub>** | 9.2 (8.6-13.0) | 21.8 (16.6-25.6)* | 7.5 (6.6-9.1) | 11.8 (10.5-15.2)* |
| **HR** | 101.4 +/-15.2 | 130.7 +/-19.1* | 98.1 +/-15.2 | 116.3 +/-15.9* |
| **SV** | 44.5 (42.6-49.3) | 51.0 (41.3-53.3) | 44.2 (42.1-48.8) | 44.6 (42.0-50.5) |
| **CO** | 4.7 +/-1.3 | 6.4 +/-1.5* | 4.5 +/-1.3 | 5.3 +/-0.9 |
| **ΔavO<sub>2</sub>** | 8.7 +/-1.9 | 12.9 +/-2.5* | 7.2 +/-2.7 | 9.3 +/-2.5 |
| **LVEF** | 72.0 +/-4.5 | 75.3 +/-3.8 | 67.9 +/-5.1 | 73.8 +/-5.8* |
| **LVEDV** | 67.5 (63.3-75.4) | 64.4 (60.2-74.4) | 71.7 (66.7-78.6) | 73.0 (63.9-77.8) |
| **LVESV** | 17.9 (15.7-21.6) | 15.8 (13.2-18.0) | 22.4 (18.4-26.0) | 17.7 (15.8-21.9) |
| **RVEF** | 63.5 (53.0-65.5) | 70.5 (66.0-73.8)* | 61.5 (52.0-68.5) | 74.0 (67.2-75.0)* |
| **RVEDV** | 85.6 (75.0-111.0) | 65.2 (63.5-78.8) | 88.6 (79.8-113.0) | 72.3 (64.0-81.4)* |
| **RVESV** | 32.5 (25.6-52.1) | 21.0 (18.4-22.3)* | 34.5 (28.6-44.5) | 18.3 (16.3-27.7)* |

PT - patients, HC - healthy controls, VO<sub>2</sub> - oxygen consumption, HR - heart rate, SV - stroke volume, CO - cardiac output, ΔavO<sub>2</sub> - tissue oxygen extraction, LVEF - left ventricular ejection
fraction, LVEDV - left ventricular end-diastolic volume, LVESV - left ventricular end-systolic volume, RVEF - right ventricular ejection fraction, RVEDV - right ventricular end-diastolic volume, RVESV - right ventricular end-systolic volume, p-value – significant differences between patients and healthy controls

Whilst exercising on the conventional ergometer, healthy controls had significantly higher peak CO (P=0.02) and ΔavO_2_ (P=0.0014) than patients. However, there were no differences in peak CO (P=0.22) and ΔavO_2_ (P=0.096) between patients and healthy controls using the novel device (Figure 4B). This was despite both CO and ΔavO_2_ increasing significantly in patients and controls groups using both exercise devices (P<0.001).

Peak RVEF was significantly lower, and peak RVESV significantly higher, in patients compared to controls using both devices (P≤0.02). This was because RVESV decreased significantly at peak exercise in controls using both devices (P≤0.002), but not in patients (conventional P=0.86; novel P=0.67, Figure 4C). RVEDV did not significantly change in patients or controls using the novel apparatus but did fall significantly (P=0.002) in controls using the conventional ergometer.

Peak LVEF was significantly lower in patients compared to controls using the novel exercise device, but not with the conventional ergometer (novel P=0.029; conventional P=0.20, Figure 4D). Peak LVEDV and LVESV were not different between patients and controls with either exercise devices.

## Discussion

The main findings of this study were: i) exercise protocols using our novel device produced significant changes in CMR-CPET metrics, ii) there was moderate correlation in exercise CMR-CPET metrics (peak VO_2_, peak HR, peak SV, peak CO, peak ΔavO_2_, and peak ventricular volumes) evaluated using the novel device and conventional MR-compatible ergometer and iii) differences between healthy volunteers and patients using both techniques were significant for most CMR-CPET metrics.

Exercise CMR techniques have gained increased attention in clinical practice for a wide range of purposes. Several studies have shown the utility of measuring biventricular function during exercise in various cardiovascular diseases (1, 12) and this is particularly pertinent to diseases like PH. There are currently two commercially available in-bore MR-compatible exercise devices that are used in most studies: the MRI cardiac ergometer (Lode BV, Groningen, The Netherlands), and the cardio step module (Ergospect, Innsbruck, Austria) (2, 10, 13, 14).

Unfortunately, commercially available devices can cost in excess of $40k and are often difficult to use because of their large size and lack of portability. In fact, compared to our novel device that takes less than a minute to set-up, a conventional MR-ergometer can take up to 10 minutes. Lower cost non-commercial alternatives do exist and include MR-compatible cycle ergometers and stepper apparatus (8, 9), but they have relatively complex designs that limit dissemination and ease of use. We have taken a slightly different approach that aims to significantly simplify both the device and the exercise protocol to keep costs low, reduce set-up time, and aid dissemination to other centers. Our low cost (∼$50), highly portable solution can easily be manufactured with components available in most hardware stores. Importantly, we have also included detailed schematics, as well as the ramped bleep audio files, allowing our device and protocol to be simply replicated. The ease of set-up and relatively short protocol means that our approach could easily be added to the end of a routine CMR scan, adding around 10 minutes.

A vital requirement of introducing a novel MR-compatible exercise device is comprehensive comparison with the current reference standard, which in this case is a commercial MR-compatible ergometer. We demonstrated moderate correlation for most CMR-CPET metrics between the conventional and novel exercise device. Nevertheless, peak VO_2_, CO, and ΔavO_2_ were systematically lower using our novel device. This was most likely due to two reasons: i) the peak work on the novel device was only related to the frequency of motion rather than the resistance and this potentially created a ceiling affect, and ii) less muscles being used whilst exercising on the novel device, which is also the for the similar differences seen between treadmill and cycle testing (15). In addition, it should be noted that the moderate correlation between the two devices was primarily driven by the greater range of metrics in patients. However, as exercise testing is more important in patients than healthy controls, we believe that this moderate correlation implies that our novel device may have some clinical utility. Regarding clinical utility, a more central consideration is whether conventional and novel devices elicit similar differences between heathy controls and patients.

We demonstrated that our novel device was able to elicit the expected differences in CMR-CPET metrics between healthy controls and patients with PH, in whom exercise intolerance and RV dysfunction is well documented (2, 16, 17). We were able to show that both the conventional and novel devices were able to elicit significant differences in peak VO_2_, HR, RVEF and RVESV between PAH patients and healthy controls. Furthermore, using our novel device we were able to demonstrate a difference in peak LVEF between PAH patients and healthy controls, which was not elicited with the conventional bike device. This is an interesting finding as many of our patients also had systemic sclerosis, and we believe our novel exercise protocol was able to unmask underlying LV disease (18). The reason that the conventional bike does not unmask LV dysfunction is not obvious but could result from differences in the mechanics of the exercise performed that might affect venous return or afterload. Conversely, we also showed that our novel device was not able to show significant differences between patients and healthy controls in CO and ΔavO_2_. These findings suggest that our novel device would be less suited to combined CMR-CPET studies that aim to elicit differences in tissue oxygen extraction. Nonetheless, we do believe that our cheap and easy to use device would be sufficient to demonstrate differences in contractile reserve.

## Limitations

The main limitation of this study is that the amount of work performed on our device was only associated with the frequency of motion. This means that there is a significant ceiling to the maximum work achievable on our novel device (because the maximum leg frequency is limited in most subjects). Conventional ergometers increase work though increasing resistance, and this may elicit a more physiological response. Nevertheless, we have shown significant increases in ejection fraction and cardiac output, demonstrating that our method is an adequate stressor in PAH patients. A further limitation was that we only investigated a single exercise ramp protocol, empirically chosen for the novel exercise device. It is possible that a different protocol might elicit different physiological responses that more closely mirror conventional ergometry. However, to test more than one protocol would not have been feasible, requiring more than two studies to be performed on each participant. This is also the reason that we could not evaluate test re-test, as this would have required 4 exercise scans to be performed on each subject. Nevertheless, with this protocol we have shown that our novel device is able to elicit differences between patients and controls in peak exercise values, particularly related to contractile reserve. Future studies should investigate protocols that could better evaluate aspects of exercise such as tissue oxygen extraction, as well as myocardial perfusion and regional wall motion abnormalities. In addition, more robust evaluation of reproducibility is necessary before any clinical use. A final significant limitation of our study was that we only investigated the differences in between healthy controls and patients with PAH. Thus, we are unable to definitively extend our findings to other important cardiovascular diseases including left ventricular failure and most importantly ischaemic heart disease. Thus, future studies should focus on diversifying patient populations and potential stress applications. It should also be noted that our design relies on the specific configuration of our scanner that allows the base to fit into a space usually occupied by a cushion. This space is not present on all scanners, and we have therefore included an alternative fixation method based on Velcro fastenings (Supplementary Figure 2 and Video 2). This method should be applicable to most scanners systems and is still low cost and simple.

## Conclusions

We have created a simple, easy to use and affordable exercise apparatus for CMR environment. Peak exercise metrics correlate reasonably well with those achieved using conventional MR-compatible supine ergometry. Given its portability and low-cost to build, this may encourage greater dissemination of exercise CMR in clinical and research practice.

## Supporting information

Metronome file

## Data Availability

All data are incorporated into the article and its online supplementary material. We do not have local ethical approval to make the study dataset publicly available. However, the study dataset will be made available to other researchers for purposes of reproducing the results or replicating the procedure upon reasonable request to the corresponding author, subject to institutional and ethical committee approvals.

