## Supplementary material for "Validation of a novel, low-cost, portable MRI-compatible exercise device in healthy volunteers and patients with pulmonary hypertension"

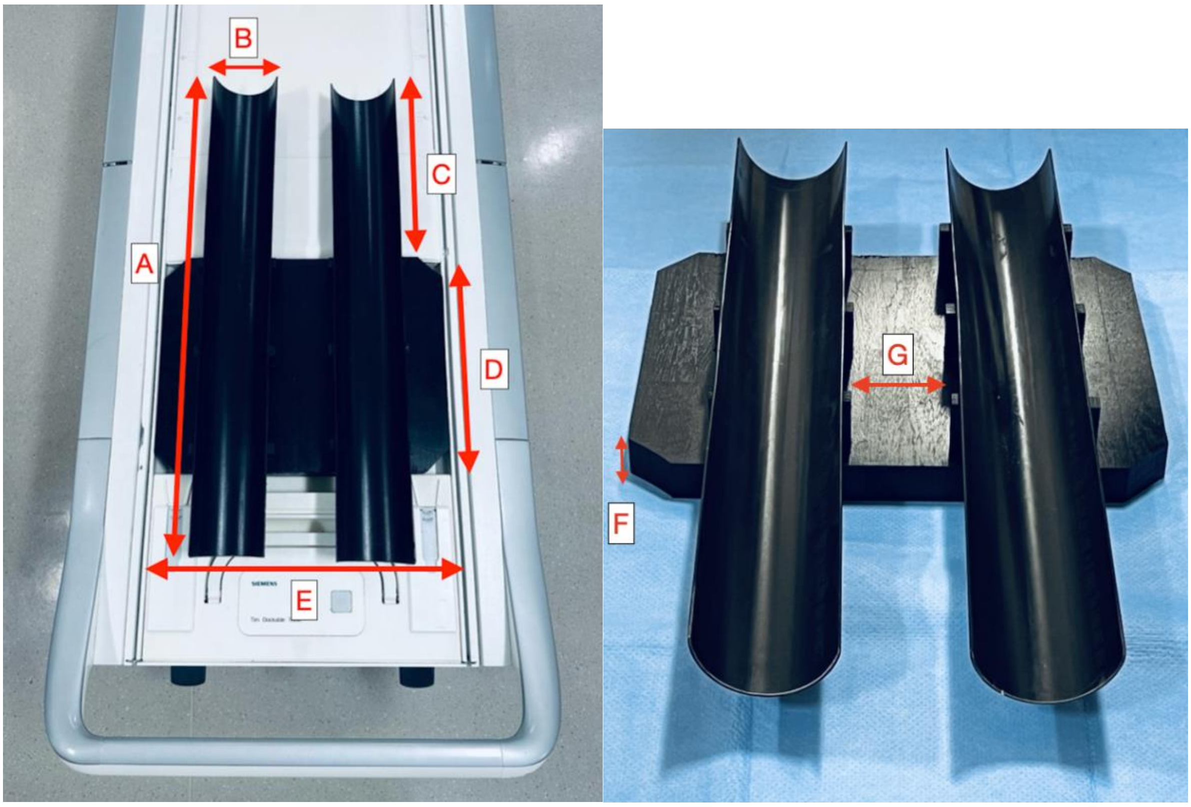


Supplementary Figure 1. Measurements of the novel exercise apparatus. A - 83.3 cm, B- 11.2 cm, C- 31.5 cm, D- 38 cm, E- 48.4 cm, F- 3.8 cm, G- 10.5 cm

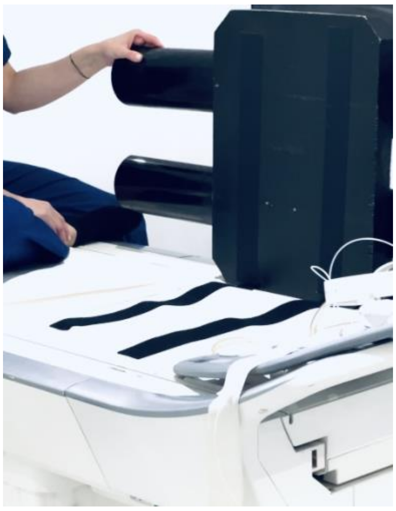

Supplementary Figure 2. Heavy Duty Velcro Attachments for fixating the device to the bed.

Supplementary video 1. Healthy control exercising with the novel exercise apparatus moving their legs back and forth in time with a metronome of gradually increasing speed.

Supplementary audio file. 18 minute file with bleeps increasing in frequency as shown below.

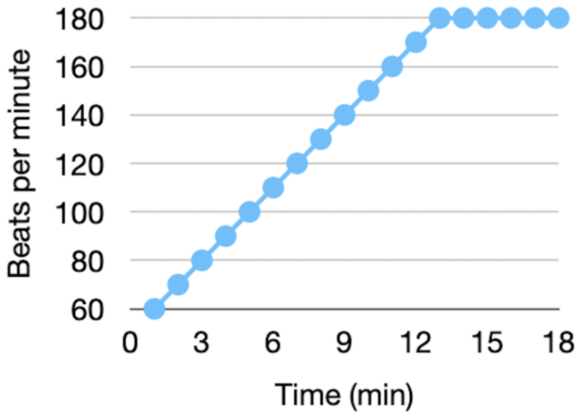
180
160
140
120
100
80
60
Beats per minute
0
3
6
9
12
15
18
Time (min)
